# Development and Validation of a Rapid Health Literacy Assessment Tool for Chinese Primary and Secondary School Students

**DOI:** 10.1101/2025.08.12.25331771

**Authors:** Jiayue Guo, Ziqing Zan, Lizhu Liu, Yuxing Wang, Mengyu Li, Lili You

## Abstract

Health literacy (HL) plays a critical role in shaping health behaviors in children and adolescents. Although children and adolescents are marked by increased involvement in health decision-making, most HL studies and measures of HL have focused primarily on adults. There is a lack of an appropriate test-based validated tool for measuring comprehensive health literacy. This study aimed to develop a rapid-assessment health literacy questionnaire for primary and secondary students to address gaps in existing tools. A health literacy indicator system was first established through literature analysis, the Delphi method, and expert interviews. Items were then generated for each domain of the indicator system. Finally, a stratified sampling survey with 3,325 primary school students and 2,788 secondary school students was conducted. The Rasch model was applied to evaluate reliability and validity. Reliability analysis demonstrated favorable overall fit between observed data and the model. Validity results showed internal consistency. We developed a robust and contextually tailored assessment tool for Chinese primary and secondary students, which is expected to comprehensively measure HL of students aged 6 to 15 years. Future cross-cultural validation is recommended to enhance its global applicability.

**Contribution to Health Promotion:**

- Health literacy is a social determinant of health. Existing tools for children and adolescents vary in approach and design, but few have focused on comprehensive health literacy encompassing broad health domains such as hygiene, physical activity, nutrition, mental health, e-health, safety, and disease prevention.
- This paper describes the development and validation of the Chinese Rapid Health Literacy Questionnaire for primary and secondary students (CRHLQ-PS), an innovative, comprehensive tool tailored to children and adolescents.
- The CRHLQ-PS can support the development of health literacy–enhancing interventions by capturing multiple dimensions of health literacy across health promotion settings.

## Introduction

Health literacy (HL) has been defined as “the integration of skills, knowledge, and motivational drivers to access, understand, appraise, and apply health information for decision-making in healthcare, disease prevention, and health promotion contexts”(Liu et al. 2020; Sørensen et al. 2012). The WHO Integrated Model(Santana et al. 2021) synthesizes individual and population perspectives, highlighting HL as a dynamic process that requires alignment with societal systems to achieve health equity. Childhood and adolescence are formative periods for acquiring health behaviors, and HL promotion during this stages can significantly impact the quality of life in adulthood(Fleary, Joseph, and Pappagianopoulos 2018). Children’s and adolescents’ HL is multidimensional, extending beyond basic numeracy and literacy skills, and developmental characteristics is related to the application of HL skills(Joseph and Fleary 2021; Massey et al. 2012). The WHO has undertaken various initiatives to promote HL as an enabling factor for health promotion within a whole-of-society approach(World Health Organization 2017).

Current studies have developed some domain-specific assessment tools for children’s and adolescents’ HL, including nutritional/food literacy(Kelly and Nash 2021; Truman, Lane, and Elliott 2017), physical health literacy(Edwards et al. 2017), and mental health literacy(Bhagat, Howard, and Aldoory 2018; Clark et al. 2020). However, there remains a lack of an appropriate tool for measuring comprehensive children’ and adolescents’ HL—one that employs grounded theory to identify key HL attributes while recognizing it as a latent construct. By covering multiple health domains, such a tool can capture the common daily activities in which children engage in health-promoting behaviors, ensuring that HL tool development encompasses all relevant dimensions(Van Boxtel et al. 2024).

Existing HL measurements have some main deficiencies: (1) insufficient cultural sensitivity: many tools failed to fully capture cultural factors(Spillane et al. 2020); (2) insufficient psychometric properties and quality lacking(Haun et al. 2014), which represents a severe weakness in a measure termed construct under-representation(Hawkins, Elsworth, and Osborne 2018). (3) poor age adaptability: many measures are adapted from adult-centric instruments such as NVS(Caldwell and Killingsworth 2022) and s-TOFHLA(Chang, Hsieh, and Liu 2012) or lacking age-span perspective(Chou, Liao, and Chen 2023), resulting in less precise evaluation across different age groups. (4) insufficient content coverage: tools often emphasize functional health literacy while neglecting the assessment of interactive and critical health literacy. (5) absence of digital health literacy. With the popularity of digital media, digital health literacy has become increasingly important(Arias López et al. 2023; Ban, Kim, and Seomun 2024), yet some tools fail to fully assess the capabilities in this regard(Chou et al. 2023). Furthermore, studies examining frameworks as a guide for developing HL measures for children and adolescents indicate theoretical insufficiency and fragmentation. HL conceptualization has largely been largely limited to adult HL in healthcare and disease prevention contexts(Bröder et al. 2017; Guo et al. 2018; Okan et al. 2015; Ormshaw, Paakkari, and Kannas 2013), and few tools have involved children and adolescents in their development(Fretian et al. 2020). In addition, varying definitions and instruments across studies make comparisons and integration of results difficult(Urstad et al. 2022).

The challenge is to develop a more comprehensive, highly adaptable, and fully validated HL questionnaire that is context- and content-specific, aimed at better supporting health education and intervention efforts for children and adolescents. To achieve this, it is essential to develop a tool sensitive to the diverse HL needs of children and adolescents, capturing key mechanisms that can inform intervention design and evaluate intervention effects. From a health promotion perspective, stage- and developmentally-targeted HL modeling is therefore vital. In particular, we fully consider the increasing independence of children and adolescents in areas such as food choices, physical activity, sleep patterns, and sexual expression throughout the transitional years.

Previous research has often relied on classical test theory (CTT) and item response theory (IRT) to validate scale reliability(Mansfield, Patalay, and Humphrey 2020). The CTT faces limitations with measurement and sample dependency, whereas IRT correlates examinee ability with the likelihood of correct responses. Rasch analysis provides a probabilistic approach for scale scoring, allowing both respondent ability and item difficulty to be quantified on a single, unified scale(Jabrayilov, Emons, and Sijtsma 2016; Woudstra et al. 2019). Therefore, we employed Rasch analysis to evaluate the questionnaire’s difficulty, accuracy, and psychometric properties.

In response to previous research findings, this study aims to conceptualize health literacy among children and adolescents and to develop a Chinese Rapid Health Literacy Questionnaire for primary and secondary students (CRHLQ-PS). The specific research questions are as follows: (1) How can existing HL models be adapted to account for the cognitive, social, and emotional developmental needs of children and adolescents? (2) Which domains and competencies should be prioritized in a context-specific HL assessment tool? (3) Does the proposed questionnaire demonstrate reliability and validity across different age groups?

## Methods

### Stage I: Development of the CRHLQ-PS

#### Step 1: Concept modelling

To identify candidate theories for health literacy modeling, we conducted a systematic search of PubMed, Web of Science, and CNKI covering the period 2000–2024. The search terms included “health literacy,” “eHealth literacy,” “health competence,” “health capability,” “health empowerment,” and “child/adolescent/school-age/youth” (Figure 1).

**Figure 1.**
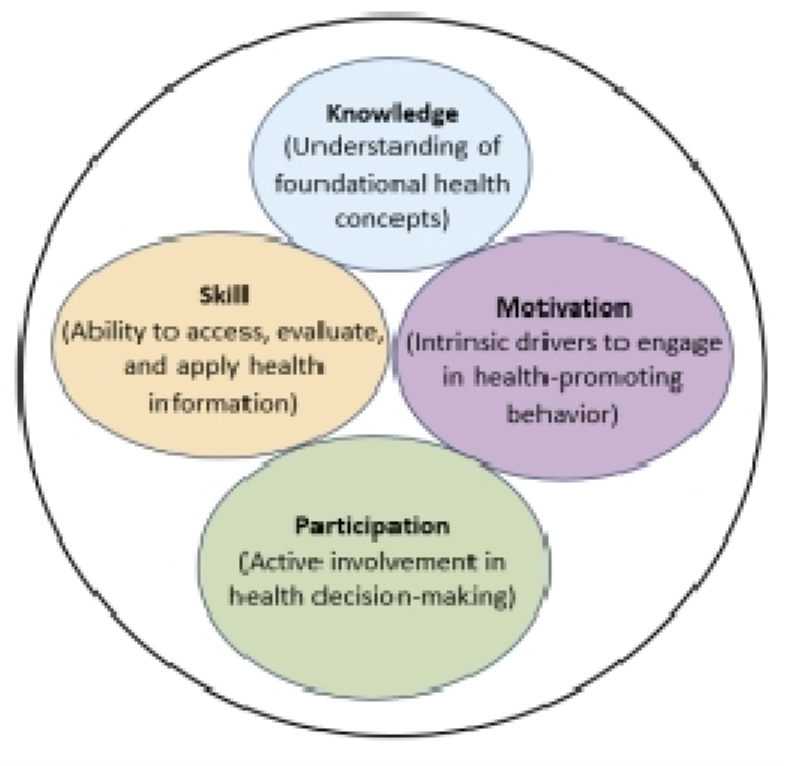
Dimensions of the health literacy conceptual model

**Figure 2.**
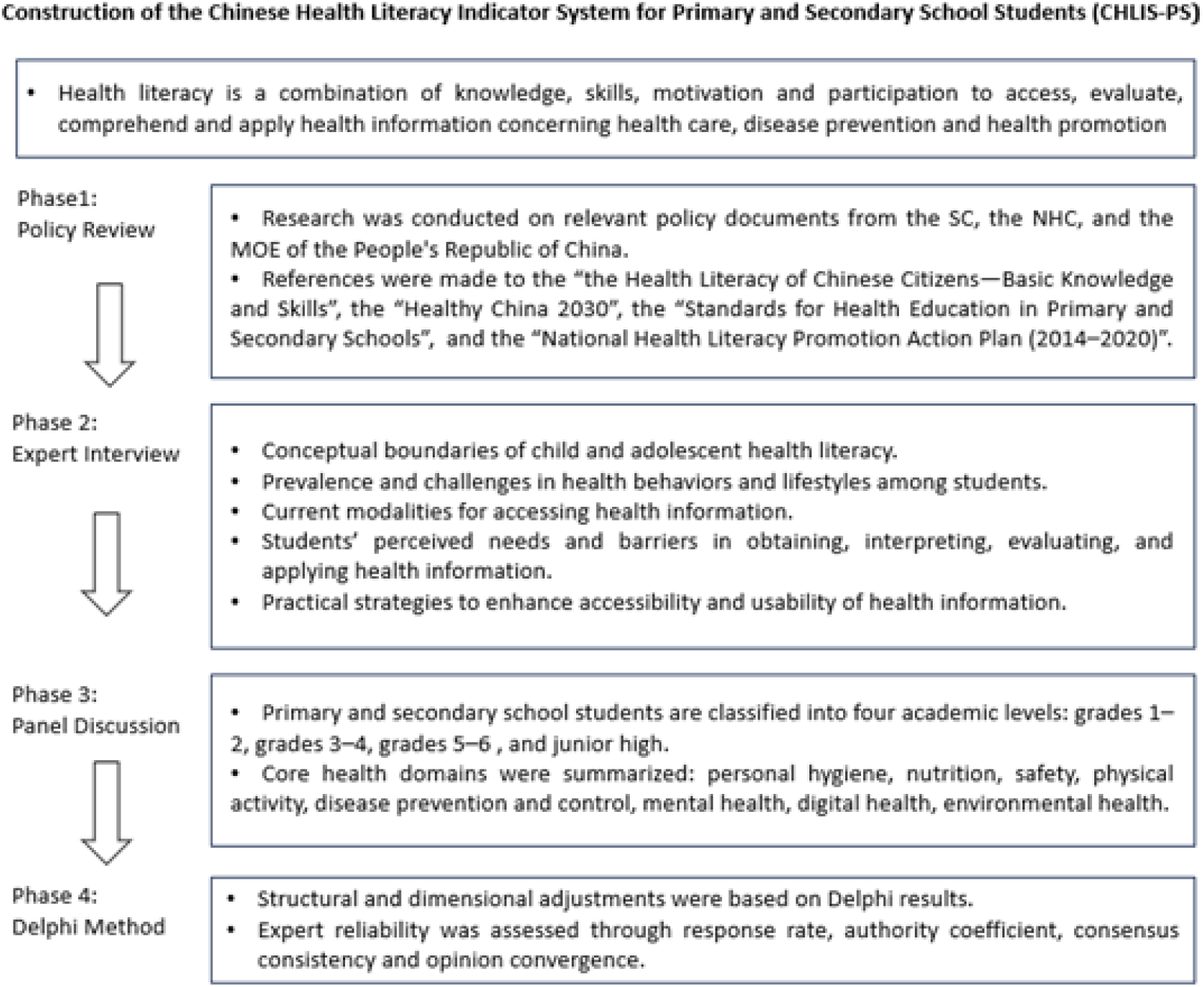
Diagram for the procedures followed to develop the health literacy indicator system

Based on Nutbeam’s hierarchical model and Bandura’s Social Cognitive Theory (SCT), we identified four core dimensions of health literacy: (1) Knowledge: understanding fundamental health concepts; (2) Skills: the ability to access, appraise, and apply health information; (3) Motivation: intrinsic drivers that promote engagement in health-enhancing behaviors; and (4) Participation: active involvement in both individual and collective health-related decision-making (Figure 1).

To address the limitations of previous adult-centric HL models, we introduced two distinctive features:(1) Developmental adaptation: acknowledging cognitive and behavioral progression, the model incorporates a developmental perspective across educational stages — early primary (grades 1–2, ages 6–8), middle primary (grades 3–4, ages 9–10), upper primary (grades 5–6, ages 11-12), and junior high (ages 13–15). (2) Domain-specific articulation: while rretaining HL as a cross-cutting capacity, the model integrates context-specific literacies validated in prior studies, including eHealth literacy, nutrition literacy, physical literacy and mental health literacy.

In our study, the identified child- and adolescent-centric HL model integrates four core competencies—knowledge, skills, motivation, and participation. Distinct from adult-centric frameworks, it advances HL measurement and intervention by incorporating developmental adaptation and domain-specific articulation, ensuring accuracy across evolving capacities and real-world health contexts.

#### Step 2: Indicator construction

##### Phase 1

*Policy Review:* Firstly, we collected and analyzed policy documents relevant to HL from Chinese government’ websites, including the State Council of the People’s Republic of China (SC), the National Health Commission of the People’s Republic of China (NHC), and the Ministry of Education of the People’s Republic of China (MOE) (Supplementary S1-Table 1).

**Table 1.** The importance and appropriateness ratings in the Delphi method.

| Domain Indicator | Aspect Indicator | Importance |  | Appropriateness |  |
| --- | --- | --- | --- | --- | --- |
|  |  | x±s, score | CV | x±s, score | CV |
| Hygiene Literacy |  | 4.5±0.8 | 0.17 | 4.6±0.7 | 0.16 |
|  | eye health | 4.8±0.6 | 0.13 | 4.9±0.3 | 0.05 |
|  | oral health | 4.8±0.6 | 0.13 | 4.8±0.6 | 0.12 |
|  | hand health | 4.1±1.4 | 0.34 | 4.3±1.2 | 0.27 |
|  | sexual and reproductive hygiene | 4.9±0.3 | 0.06 | 5.0±0.0 | 0.00 |
| Nutrition Literacy |  | 4.8±0.8 | 0.17 | 4.6±0.8 | 0.18 |
|  | nutrition | 4.9±0.3 | 0.06 | 4.8±0.5 | 0.11 |
|  | food sanitation | 4.9±0.3 | 0.06 | 4.8±0.5 | 0.11 |
| Physical Activity |  | 5.0±0.0 | 0.00 | 4.9±0.3 | 0.05 |
| Literacy | exercise | 4.7±1.1 | 0.24 | 5.0±0.0 | 0.00 |
|  | sedentary behavior | 4.8±0.4 | 0.08 | 4.8±0.5 | 0.11 |
|  | sleep | 5.0±0.0 | 0.00 | 4.9±0.3 | 0.05 |
| Preventive Health |  | 4.9±0.3 | 0.06 | 4.9±0.3 | 0.05 |
| Literacy | nutrition-related disease | 4.5±0.8 | 0.17 | 4.5±0.8 | 0.17 |
|  | infectious disease | 4.9±0.3 | 0.06 | 4.9±0.3 | 0.06 |
|  | chronic disease | 3.9±1.5 | 0.38 | 4.1±1.3 | 0.32 |
| Safety Literacy |  | 4.8±0.4 | 0.08 | 5.0±0.0 | 0.00 |
|  | accidental injury prevention | 5.0±0.0 | 0.00 | 5.0±0.0 | 0.00 |
|  | natural disaster evacuation | 4.9±0.3 | 0.06 | 4.9±0.3 | 0.06 |
| Psychosocial Adaptation |  | 5.0±0.0 | 0.00 | 4.9±0.3 | 0.05 |
| Literacy | sociocultural adaptation | 5.0±0.0 | 0.00 | 5.0±0.0 | 0.00 |
|  | behavior regulation | 5.0±0.0 | 0.00 | 4.8±0.4 | 0.08 |
|  | psychological assistance | 4.8±0.6 | 0.13 | 4.5±0.8 | 0.17 |
| Digital Health Literacy |  | 4.8±0.4 | 0.08 | 4.7±0.6 | 0.13 |
|  | access | 4.7±0.6 | 0.13 | 4.8±0.6 | 0.13 |
|  | evaluation | 4.5±1.2 | 0.27 | 4.6±0.8 | 0.17 |
|  | application | 4.5±1.2 | 0.27 | 4.5±0.8 | 0.17 |

Key documents, including *Health Literacy of Chinese Citizens—Basic Knowledge and Skills*(National Health Commission (2008) n.d.) and *Healthy China 2030* (The State Council of the People’s Republic of China (2016) n.d.), emphasized the four core competencies: knowledge, skills, motivation, and participation. Developmental benchmarks from the *Standards for Health Education* guided the design of age-specific items, while the *Ecological Environment and Health Literacy (2018)* provided guidance for contextually grounded domains, such as environmental health.

##### Phase 2

*Expert Interview*: Secondly, we conducted semi-structured interviews and panel discussions with 13 multidisciplinary experts to explore key themes, including: (1) the conceptual boundaries of children’s and adolescents’ HL; (2) the challenges in promoting students’ health behaviors and lifestyles; (3) current modalities for accessing health information (e.g., digital platforms, school-based resources); (4) students’ perceived needs and barriers in obtaining, interpreting, evaluating, and applying health information; and (5) Practical strategies to enhance accessibility and usability of health information.

##### Phase 3

*Panel Discussion*: Third, eight health domain indicators were identified through multiple brainstorming sessions and group discussions. These domains encompass several key aspects, including eye, oral, and hand hygiene; food safety and nutrition; exercise, sedentary behavior and sleep; injury prevention and natural disaster evacuation; nutrition-related, infectious, chronic and endemic diseases; sociocultural adaptation, behavior regulation and psychological support; digital health access, evaluation and application; and pollution exposure prevention and sustainable resource utilization. After phases 1,2, and 3. The initial Chinese Health Literacy Indicator System for primary and secondary students (CHLIS-PS) was developed.

##### Phase 4

*Delphi Method:* Finally, following established procedures of previous studies (Bale, Grové, and Costello 2018), the Delphi method was employed to refine the CHLIS-PS. Thirteen experts, all with expertise in health promotion, behavioral science and children health, with a minimum of 10 years of professional experience, participated in an online consultation using a structured questionnaire and proposed any additional amendments to the system. Subsequently, experts were provided with relevant background information and asked to rate each indicator in terms of importance and appropriateness (Holey et al. 2007). Expert reliability was assessed through: (1) response rate; (2) authority coefficient (Cr=[Ca+Cs]/2), where Ca (0.3-0.1) reflected judgment basis and Cs (0.2-1.0) indicated familiarity; (3) consensus consistency (Kendall’s W, 0-1); (4) opinion convergence. Open-ended feedback was used to inform iterative revision.

After repeated deliberation and consideration, “environmental health” and “endemic disease” were removed, while “sexual and reproductive health” was added. Compared with grades 5-6 and junior high school students, grades 1-2 and 3-4 did not include the domain indicator “digital health literacy”, nor the aspect indicators “chronic disease” and “sexual and reproductive hygiene” (Figure 3).

**Figure 3.**
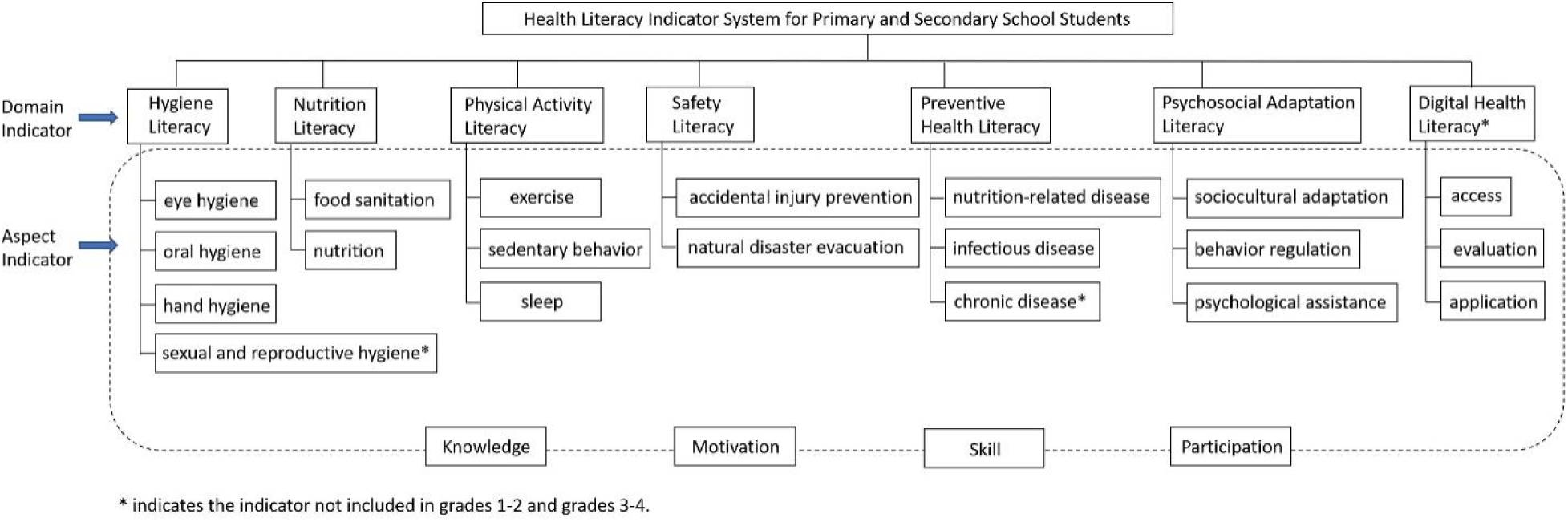
Final version of the Chinese Health Literacy Indicator System for primary and secondary students

The Delphi process yielded an authority coefficient (Cr) of 0.88, exceeding the 0.70 reliability benchmark, with Kendall’s W=0.36 (p<0.01), indicating moderate consensus. The mean importance scores for all aspect indicators ranged from 3.9 to 5.0, while the mean appropriateness scores ranged from 4.1 to 5.0 (Table 1).

#### Step 3: Item generation

We generated an item pool through three brainstorming sessions, which were subsequently merged with items independently generated by an expert. Each session lasted from 2-4 hours and was facilitated by LY. All participating researchers and experts were either health education practitioners, or health promotion specialists. For each domain indicator, we aimed to generate a minimum of four items. This approach aligns with prior findings that at least three to four items per scale are sufficient to achieve adequate internal consistency reliability(Harvey, Billings, and Nilan 1985).

Key priorities during item generation included, (1) balanced representation across each HL subdomain, (2) developmental appropriateness for children and adolescents, and (3) brevity to secure feasibility in completing the scale; consensus was reached through discussion. (Supplementary S1-Table 2)

**Table 2.**
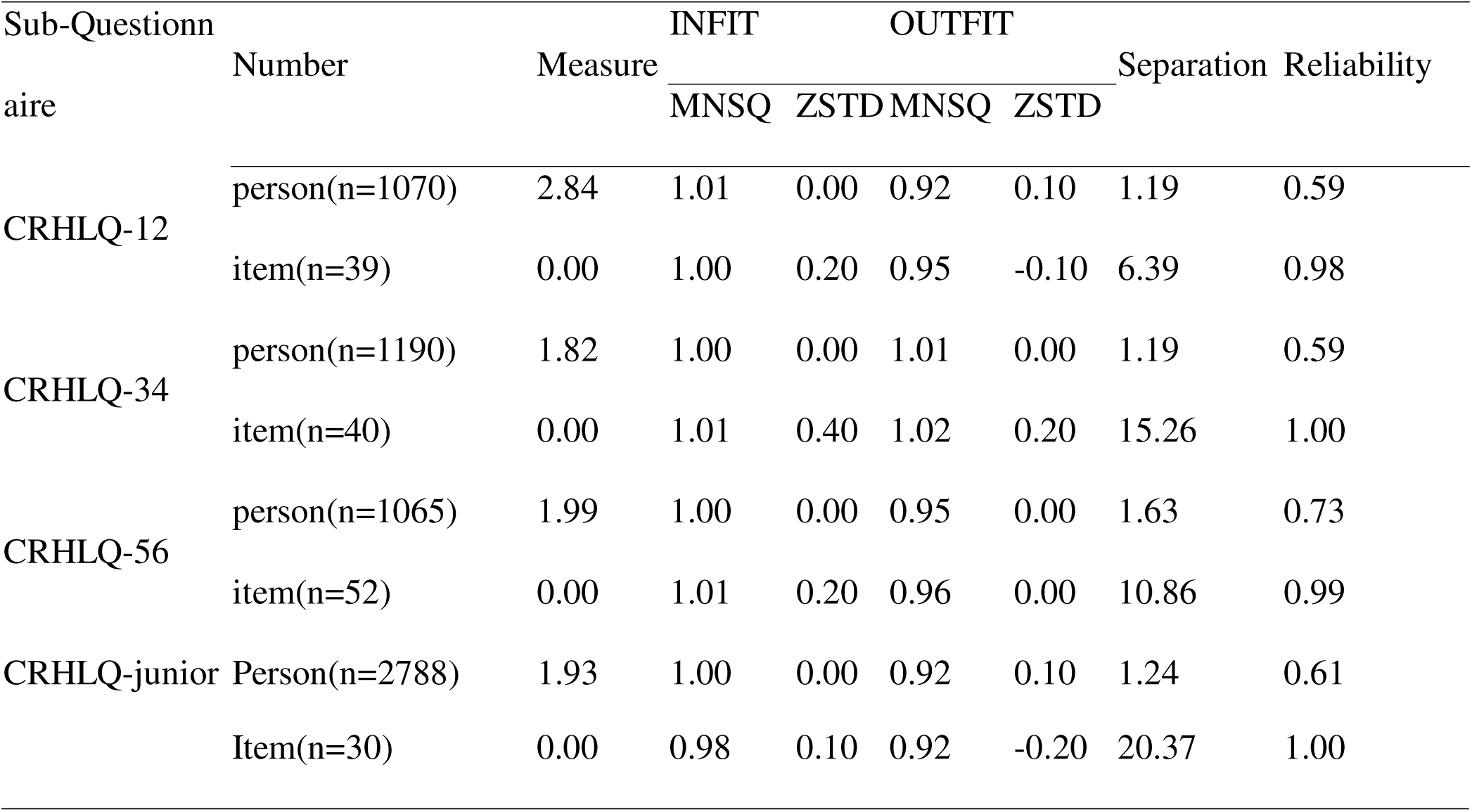
Item Summary Statistics.

#### Step 4: Questionnaire measurement

The constructed CRHLQ-PS comprises four sub-questionnaires, each tailored to specific grade levels: grades1-2 (CRHLQ-12), grades3-4 (CRHLQ-34), grades 5-6 (CRHLQ-56) and junior high (CRHLQ-junior). CRHLQ-12 contained 39 items, CRHLQ-34 contained 40 items, CRHLQ-56 contained 52 items and CRHLQ-junior contained 58 items. Question type vary across grade levels. (Supplementary S1-Table 3).

### Stage II: Validation of the CRHLQ-PS

In this phase, the developed questionnaire was administered to primary and secondary school students to evaluate its validity and reliability.

#### Data collection

A multi-site validation study using stratified sampling was conducted with 3,325 primary students and 2,788 secondary students across geographically diver regions in China via the digital platform “Yiqixiu”. Standardized administration was supervised by trained teachers, with parental assistance provided for younger students.

Ethical approval was obtained from the Institutional Review Board of the Chinese Academy of Medical Sciences & Peking Union Medical College, which oversees research involving human participants. Informed consent was obtained from the parents of all participants, following a thorough explanation of the survey’s content and objectives.

#### Statistical analysis

For CRHLQ-junior, the knowledge and skills dimensions, composed of judgmental or multiple-choice items, were analyzed using the Rasch model, while the motivation and participation dimensions, consisting of scale-type items, were evaluated through CTT analysis. The CRHLQ-12, CRHLQ-34 and CRHLQ-56 were entirely evaluated through the Rasch model.

The Rasch analysis was performed using Winsteps 3.72.3 to assess reliability and validity: reliability (>0.7) and separation indices (>2.0) confirmed measurement precision. Unidimensionality was verified through principal component analysis of residuals (first contrast eigenvalue < 2.1). Item fit was assessed through mean - square residuals (Infit/Outfit MNSQ: 0.5–1.5) and ZSTD < 2.0. The Wright map was used to calibrate the alignment between item difficulty and person ability. Model - data fit was further evaluated through Infit/Outfit MNSQ (acceptable range [0.5, 1.5]), and point - measure correlations (range 0–1] indicated item - total score correlation, with larger values indicating better discriminative power, following Rasch standards (Linacre, 2020).

Within the CTT framework, construct validity was assessed via exploratory factor analysis (EFA) with promax rotation (factor loadings > 0.4; cumulative variance ≥ 40%). Concurrent validity was examined using Spearman’s correlation analysis, and internal consistency reliability was measured by cronbach’s α (> 0.55).

## Results

### Sample characteristic

The survey included 1070 students in grades 1-2, with 792 in Luzhou, 116 in Taiyuan, 142 in Beijing, and 20 in Zhengzhou; 1190 students in grades 3-4, with 962 in Luzhou, 78 in Taiyuan, 115 in Beijing, and 35 in Zhengzhou; 1065 students in grades 5-6, with 646 in Luzhou, 162 in Taiyuan, 218 in Beijing, and 39 in Zhengzhou; and 2788 in junior high, with 1875 in Luzhou, 650 in Taiyuan, 263 in Beijing.

### Reliability analysis

For CHLQ1-12, CHLQ1-34, CHLQ1-56, and the judgmental/multiple-choice items of CRHLQ-junior, person ability values (2.84, 1.82, 1.99, 1.93) all exceeded the corresponding item difficulty value of 0. Reliability metrics, including person reliability (0.59, 0.59, 0.73, 0.61) and item reliability (0.98, 1.00, 0.99, 1.00), indicated consistent and stable measurement. Separation indices revealed person separation values (1.19, 1.19, 1.63, 1.24; below the standard 2), indicating relatively concentrated person ability levels, whereas item separation indices (6.39, 15.26, 10.86, 20.37; exceeding the ideal 2) highlighted the questionnaire’s strong discriminative power and high item difficulty differentiation. Additionally, INFIT MNSQ and OUTFIT MNSQ values for both items and persons approached the optimal value of 1, with INFIT ZSTD and OUTFIT ZSTD within the range of −2 to 2, demonstrating a good overall fit between observed data and the Rasch model (Table 2).

For scale-type question of CRHLQ-junior, the Cronbach’s α coefficients for the overall, motivation and participation dimensions were 0.818, 0.769, and 0.676, respectively.

### Validity analysis

#### Unidimensionaliy

The Rasch model measurement explained 32.1% of the total variance. The first residual factor, with a value of 2.4, accounted for 5.7% of the residual variance. These results satisfied the unidimensionality requirement, indicating that the items measured the same domain.

#### Difficulty Analysis

For CRHLQ-12, item difficulty approximated a normal distribution (mean = 0 logit), while person ability estimates also followed a roughly normal distribution, ranging across 6 logits (mean = 2.84 logit), exceeding the average item difficulty. In CRHLQ-34, most items were distributed below 0 logit, with person abilities normally distributed (mean = 2.12 logit), again higher than item difficulty. For CRHLQ-56, item difficulty approximated a normal distribution (mean = 1.99 logit); the mean person ability exceeded mean item difficulty by nearly 2.00 logits, but no items targeted high-ability students (≥2.50 logit). In CRHLQ-junior, person ability followed a normal distribution (mean = 2.12 logit), with no items corresponding to very low ability levels (below −1 logit). Overall, results showed that the overall difficulty of the questionnaire was set relatively low. The Wright maps are presented in Supplementary S2.

#### Spearman’s correlation analysis

High correlations were observed between the motivation and participation dimensions and the total score (r = 0.771 and 0.835, respectively; p < 0.001). The KMO values were 0.924 and 0.787, and Bartlett’s tests of sphericity yielded χ² = 22,586.647 and 6,668.652, respectively (p < 0.001). Principal component analysis of the motivation and participation dimensions identified two common factors with eigenvalues >1, explaining 55.21% and 51.63% of the cumulative variance, respectively. Factor loadings exceeded 0.4, communalities were >0.3, and no cross-loadings were observed, supporting good structural validity (Supplementary S1-Tables 4,5).

## Discussion

We developed and validated a rapid-assessment, comprehensive HL questionnaire, comprising four sub-questionnaires tailored to different grade levels. This study offers new insights into the conceptualization of children’s and adolescents’ HL from a developmental perspective. During questionnaire development, several key methodologies were employed. Firstly, the number and content of items were rigorously aligned with the identified indicators, while allowing for flexible adjustments to ensure contextual relevance. Second, question formats were adapted to suit different academic levels. For younger students with relatively limited comprehension skills, the questionnaire adopted simplified formats and dimension-specific measures. In contrast, 5-point Likert-type scales were employed for older students to capture a more nuanced understanding of their motivations and engagement in health behaviors. This differentiated approach enhanced both the sensitivity and accuracy of the questionnaire. Third, pictorial representations were incorporated to support younger children in understanding and responding to questions effectively.

We employed the Rasch model for quality assessment, which unifying participant ability and questionnaire difficulty on a single measurement scale(Jabrayilov et al. 2016). The results showed the questionnaire demonstrated good internal consistency, and effectively distinguished between students of varying ability levels. However, the average item difficulty was lower than the average participant ability, implying an overall simplicity. This simplicity may be attributed to the predominance of straightforward judgment-type questions. Future studies should consider incorporating more challenging question formats, such as open-ended or computational items, to increase the overall difficulty and better differentiate higher-ability respondents.

HL Improvement provides a foundational competence for health empowerment and promotes individual well-being. Previous research vary considerably on the definition and what constitutes HL has been consistently contested within the literature(Bröder et al. 2019). Recently, karolina developed the first child and adolescent-centered health literacy model(Seidl et al. 2025), highlighting the importance of directing attention to HL in younger populations. Conceptualizing HL in this context allows for recognition of empowerment that extends beyond the mere acquisition of basic health knowledge.

In this study, we constructed a robust HL indicator system for primary and secondary school students, conmprising seven domain indicators. To our knowledge, this is the first study to develop a dynamically evolving assessment framework tailored to this population.

Existing European and American systems have adapted assessment tools like s-TOFHLA(Chang et al. 2012), REALM-teen(Davis et al. 2006) and HLS-EU-Q47(Domanska et al. 2018) for children and adolescents. The s-TOFHLA REALM-teen primarily focus on text comprehension and health decision-making skills in medical scenarios, whereas HLS-EU-Q47 addresses three major domains —health promotion, disease prevention, and healthcare— emphasizing cross-cultural adaptation and the evaluation of policy interventions. Additionally. the WHO GAMA framework’s 47 adolescent health indicators (World Health Organization 2024) cover health behaviors, social determinants, and system performance; however, GAMA primarily target policy and system-level assessments rather than on specific measurement tools.

This study has some limitations. Data were collected based on voluntary responses from students. Given their age and level of cognitive development, the results may be subject to inherent biases. In addition, participants were recruited through cluster sampling, which could limit the representativeness of the sample. Nevertheless, it is important to note that this limitation is somewhat mitigated by the use of Rasch analysis, which is less dependent on sample characteristics compared to classical test theory.

In conclusion, the developed CHLIS-PS, validated through literature based, policy alignment and expert consensus, demonstrates robust and comprehensiveness. The CRHLQ-PS is expected to provide a comprehensive assessment of health literacy in students aged 6 to 15 years, addressing a critical gap in the field. Future cross-country comparative studies are anticipated to further validate the applicability of the CRHLQ-PS, which has the potential to become a key tool for advancing global adolescent health literacy.

## Conclusion

The Child and Adolescent Health Literacy Questionnaire for Primary and Secondary Schools (CRHLQ-PS) is expected to comprehensively measure health literacy of students aged 6 to 15 years, filling the gap in related research areas. Future cross-country comparative studies are anticipated to further validate its applicability, and the CRHLQ-PS has the potential to become a key tool for advancing global adolescent health literacy.

## Acknowledgements

LY was funded by the Science and Technology Innovation Project in Medicine and Health, Chinese Academy of Medical Sciences under Grant [number 2021-I2M-1-046] and the National Natural Science Foundation of China under Grant [number 71904205]. We acknowledge the support from the participating primary and secondary schools, and the digital platform *Yiqixiu* that facilitated data collection in China.

## Data availability statement

The data underlying this article will be shared on reasonable request to the corresponding author.

## Author Contributions

JG and LY conceptualized the study; LL and ZZ developed the methodology; ML, ZZ, and LL conducted the investigation; JG and YW performed the formal analysis; JG wrote the original draft; JG and YW reviewed and edited the manuscript; LY supervised the study and acquired funding; all authors reviewed and approved the final version of the manuscript.

